# Clinical features and outcomes of scorpion sting in western lowland of Eritrea: a prospective descriptive study

**DOI:** 10.1101/2024.01.02.24300701

**Authors:** Okbu Frezgi, Araia Berhane, Adiam Gebreyohannes, Ghide Ghebrewelde, Henok Tekie, Tsegezab Kiflezgi, Abdulaziz Mohamedsied, Yonas Tekie, Abel Alem, Hagos Ahmed, Tewaldemedhine Gebrejesus

## Abstract

**Background:** Scorpion envenomation is a public health problem that results in a life-threatening medical emergency in tropical and subtropical regions. Paediatric victims are more at risk to severe envenomation compared to adults.

**Objectives:** This study aimed to determine clinical features and outcome of patients hospitalized due to scorpion stings at the Tesseney Community Hospital.

**Methods:** A prospective, descriptive cross-sectional study conducted from 1^st^ June 2019 to 31^st^ May 2020 on patients hospitalized due to scorpion stings at the Tesseney Community Hospital.

**Results:** 165 scorpion sting patients were admitted during the study period. Majority were older than 15 years old (61.8%) with an approximately equal male-to-female sex ratio (0.94:1). Scorpion sting cases largely belonged to urban (57%) compared to rural areas. The black scorpion (38.8%) was the predominately identified scorpion, but in a proportion of cases the scorpion colour was undetermined (31.5%). The foot was predominate sting site (64.8%) followed by hand (31.5%). A single sting (91.5%) was more frequently presented compared to multiple stings (8.5%). Majority (94.8%) of scorpion sting cases occurred during the summer period with highest scorpion stings counts belonging to September and October. The main clinical manifestations upon presentation were localized pain (70.3%) and sweating (56.4%) with more severe symptoms exhibited amongst age groups less than 15 years old. The fatality rate (4.8%) was largely associated age groups less than 15 years old with (p = 0.006, OR: 2.845; 95% CI: 0.656-12.343).

**Conclusion:** Our study found children experienced more severe envenoming symptoms with related mortality compared to adults. This study may be a tool to identify at-risk population groups and build measures to prevent scorpion stings within the western lowlands of Eritrea.

## Introduction

Scorpion stings are a life-threatening medical emergency with frequent presentations within emergency department in many areas of the world.(1) Accurate global statistical evidence on scorpion stings are not readily available, but the literature indicates that all settings are usually affected by this medical threat.(2) In 2018, WHO reported that the true scorpion sting envenomation burden is not known because many accidents occur in villages within tropical and subtropical countries where victims likely do not seek medical attention.(3) It is estimated that 1.23 million scorpion stings occur per year with roughly 3250 deaths per year.(1,2,4–6) Developing countries are often reported with higher scorpion envenomation accidents and related mortality compared to developed countries due to lower socioeconomic structures and inadequately equipped health facilities.(2) Pediatric victims are more at risk to severe envenomation with related mortality compared to adults due to their small body size.(4,7)

Scorpions are predatory arachnids with four pairs of legs, a pair of grasping pedipalps, and a narrow-segmented tail with a characteristic curved back ending with a venomous sting.(5) Scorpions are generally located in dry and hot environments, although some species also occur in forest and wet savannahs.(5) All scorpion species are nocturnal, hiding under stones, wood, or tree bark during the day.(5,8) They are coated in a variety of colours, ranging from straw colour to yellow, and light brown to black.(4) Scorpions have a remarkable capacity to survive and adapt, allowing them to withstand heat, drought, desert and freezing conditions which permits the species to survive virtually independently of their environmental constraints.(5) Currently there are more than 1500 subspecies of scorpion recognized worldwide, with 50 subspecies having dangerous venom for humans.(4,8) The *Parabuthus* (western and southern Africa), *Hottentotta* (south Africa and south-east Asia), *Leiurus* (northern Africa and middle East), and *Androctonus* (northern Africa and southeast Asia) scorpion species are reported to be found within the African continent.(5) Within East Africa, scorpion sting cases are common in the central-west regions of Sudan due to geographical location, climate, and the socioeconomic structure.(8,9) Though there is no ecological survey of scorpion species done within Eritrea, scorpion sting accidents are found to be common in the western lowlands of Eritrea which borders Sudan.

Scorpion venom is a “cocktail” of complex structures formed by neurotoxic proteins, acidic proteins, salts, and organic compounds, with variable compositions and lethality amongst subspecies.(4,5,8) Venom mainly effects neurologic, cardiovascular, respiratory, hematologic, and renal systems with local effects around the sting site (i.e. redness, pain, swelling, and burning).(4,8) Toxins and enzymes within venom possess neurological tropism effects that act mainly on voltage-gated sodium and potassium ion channels on excitable cells of nerves and muscles.(5) Venom from the same scorpion can have multiple toxins that can interact with each other, modulating the response of the ion channels involved and leading to complex, rapidly progressive symptoms.(5–8,10–12) The main effect of the venom in a victim is to mediate ion channels for intense, persistent depolarization of autonomic nerves, leading to massive discharges of neurotransmitters from both autonomic nervous system branches.(5) This is referred to as an “autonomic storm” owing to the sudden pouring of endogenous catecholamine into the circulation.(5,11–14) The effects of an “autonomic storm” are seen throughout the body, but are more serious on the cardiovascular and respiratory systems leading to arrhythmia, hypertension or hypotension, and pulmonary edema.(15) Clinically, the “autonomic storm” evoked due to scorpion envenoming is characterized by transient parasympathetic (vomiting, profuse sweating, ropy salivation, bradycardia, ventricular premature contraction, priapism in male, and hypotension) and prolonged sympathetic (cold extremities, hypertension, tachycardia, pulmonary edema, and shock) stimulation.(5,16–19)

Signs and symptoms of scorpion stings are divided into three classes commonly known as the Abroug’s classification. (4–6,8,11,14,20) Majority of scorpion sting cases only require supportive therapy, including ibuprofen, cleaning of the sting area, and tetanus prophylaxis. In certain areas of the world where scorpions are more poisonous (i.e., the Middle East), scorpion stings are treated with drugs and methods that reduce symptoms and complications.(21) Over the course of the 20^th^ century, scorpion antivenom (SAV), vasodilators, and intensive care facilities have significantly reduced the fatality rate of severe scorpion envenomation.(5) Administering SAV can neutralize unbound circulating venom to reduce the probability of sympathetic nervous system over-stimulation.(5) Though the benefits of SAV has been proven in some studies, the effectiveness of SAV continues to be debated.(4,5,19,22) A critical component of scorpion sting pathogenesis is alpha-receptor stimulation.(5,14) Prazosin is an α_1_-blocker commonly known as a pharmacological and physiological antidote to the venom action. The drug acts to relieve serious breathing difficulty due to high blood pressure.(5,12) As a phosphodiesterase inhibitor, the drug reduces blood pressure via reducing preload and left ventricular impedance without increasing heart rate.(5) Previous studies have recommend prazosin as a first-line treatment for patient arriving to the ED with scorpion envenoming due to the drugs high availability in rural settings.(12,23)

This study aims to describe the clinical manifestations and outcomes of scorpion sting patient cases admitted at Tesseney Community Hospital. The study seeks to inform medical professionals and public health workers to design better preventive measures to reduce scorpion sting incidents within Eritrea.

## Objective

### General objective

The study aims to provide clinical features and outcomes of patient admitted to Tesseney Community Hospital due to scorpion stings.

### Specific objective

To describe the clinical features of scorpion stings and to determine the risk factors associated with fatal outcomes.

## Materials and Methods

### Study design

This was a prospective, descriptive cross-sectional study conducted from 1^st^ June 2019 to 31^st^ May 2020 for patients admitted to due to scorpion stings at the Tesseney Community Hospital.

### Study area

Tesseney hospital is a community hospital located in Gash-Barka region of Eritrea that serves a catchment population of 87,992 distributed in an area of 1,096.83 km^2^ (Figure 2). The hospital provides inpatient and outpatient services, delivery service, laboratory services, imaging unit, physiotherapy unit, and possess a 115-bed capacity.

**Figure 1.**
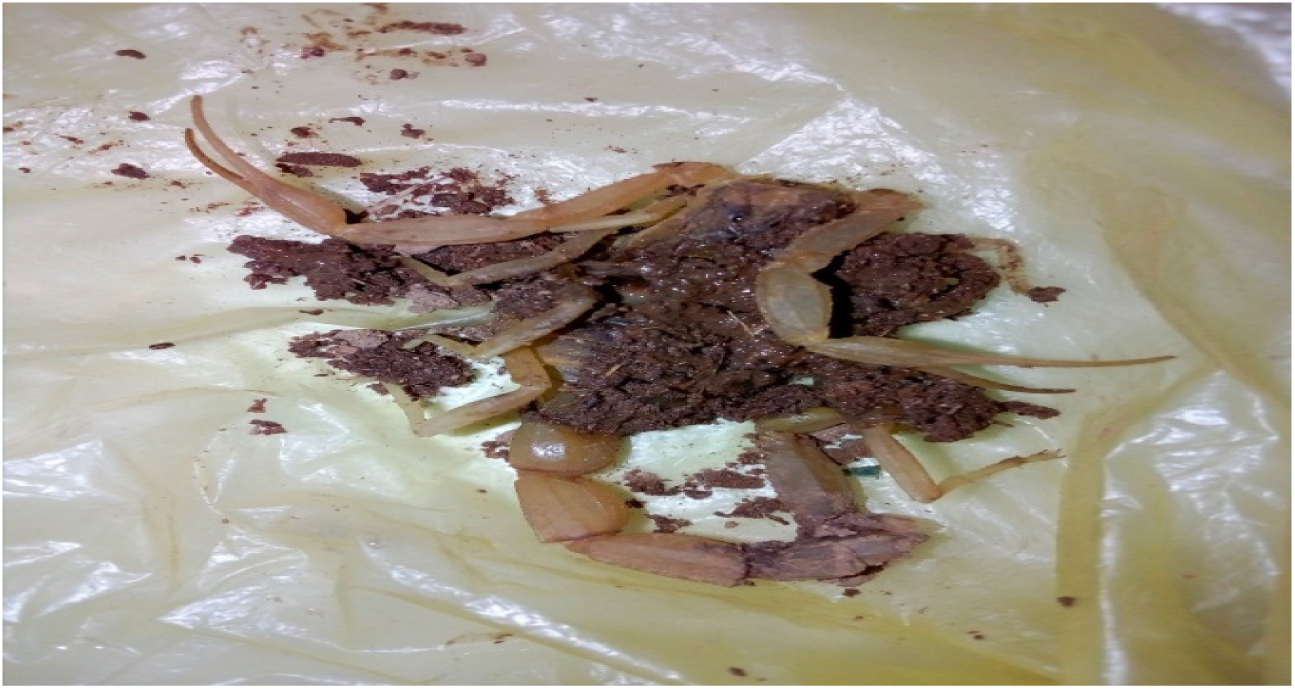
Yellow-colored scorpion brought into the ED by family members. Envenomated patient suffered from irreversible shock and pulmonary edema and was thereafter deceased.

**Figure 2.**
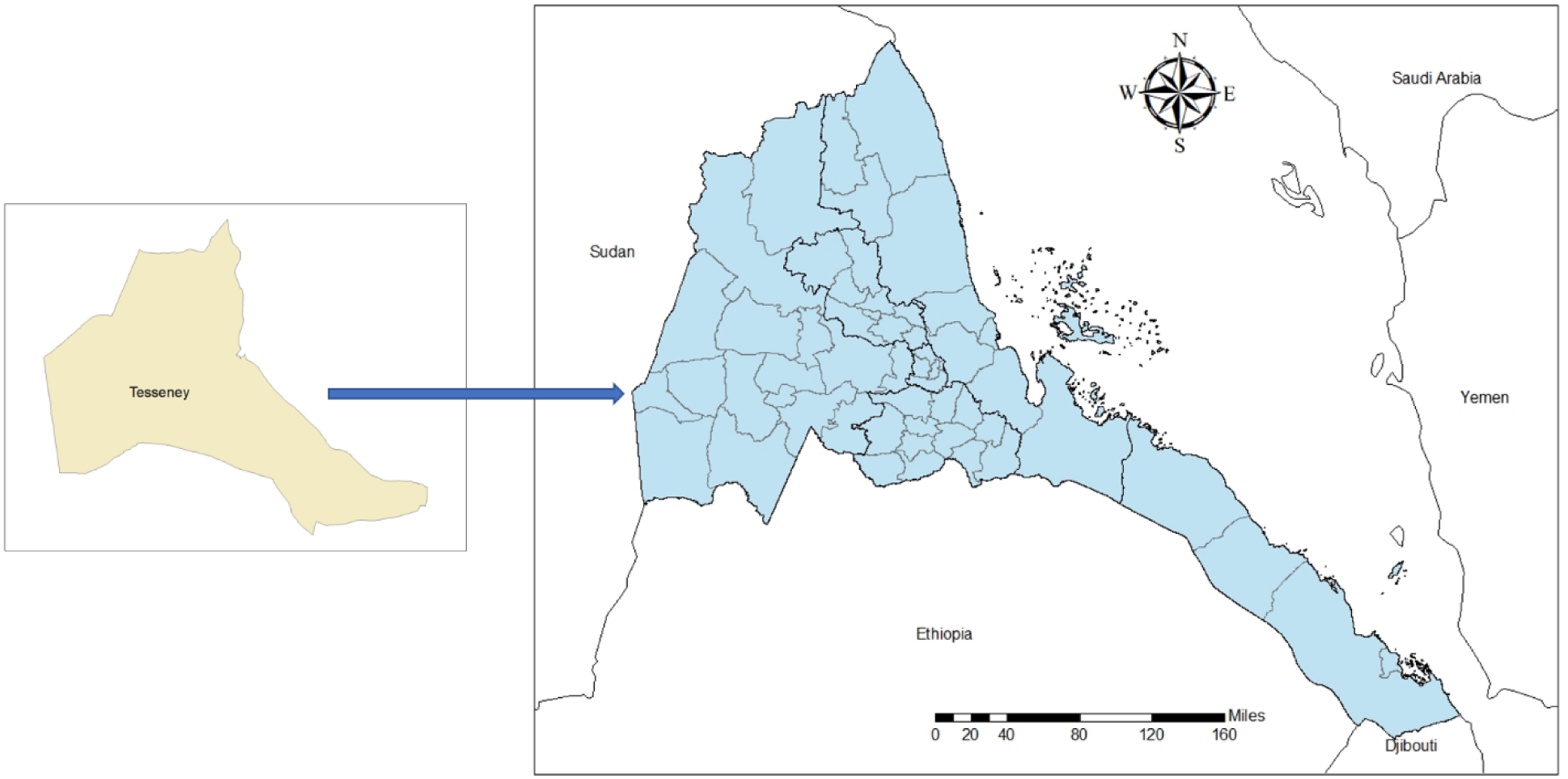
Map of Eritrea, Tesseney subzone.

### Study population

All patients admitted to the hospital due to scorpion stings from 1^st^ June 2019 to 31^st^ May 2020 were considered the study population.

### Inclusion and exclusion criteria

Patient who had a history of scorpion sting and had seen or killed the scorpion (via the victim or bystander) were also included as the study. Patients with a history of unknown bites, arrived to the ED without signs of life, and/or deceased after discharge from the hospital were excluded from the study.

### Data collection and analysis

Data was collected by admitting doctors in a predesigned questionnaire survey prepared in open data kit (ODK) and were followed until the final outcome. Demographic characteristics, vital signs, symptoms and signs, time interval between scorpion sting incident and arrival at the hospital, and medications administered were recorded. Statistical analyses were performed via Statistical Package for the Social Sciences (SPSS) v.23 software. The symptoms and signs were categorized into three classes according to Abroug’s classification (Table 1). The Abroug’s classification was utilized due to the catalogue’s convenience and its common use in neighbouring countries (Table 1).(4–6,8,11,14,20) Descriptive data were expressed as frequency and percentage while intergroup differences were analysed using Chi-Square or Fisher test depending on the characteristics of the data (p-value < 0.05 as statistically significant).

**Table 1.** Abroug’s classification.

| Classes | Signs and Symptoms |
| --- | --- |
| Class I | Pain and/or paraesthesia at the scorpion sting site |
| Class II | Fever, chills, excessive sweating, nausea, vomiting, diarrhea, hypertension and priapism. |
| Class III | Cardiovascular, respiratory, and/or neurologic |

### Ethical approval

The study received ethical approval from the Ministry of Health Research and Ethics Approval Committee. A written informed consent was obtained from each patient or parent/care taker for children. Confidentiality of study participants was maintained by analysing anonymised aggregate data and coding the personal identifiers. Patients were reassured no environmental or occupational harm by participating within this study. All patient subjects were provided the available standard treatment for their disease process.

## Results

A total of 165 patients with scorpion stings were admitted to the Tesseney community hospital from 1^st^ June 2019 to 31^st^ May 2020. Our results had an almost similar sex distribution (male = 48.5%; female = 51.5%) with a male-to-female sex ratio of 0.94:1. The majority of cases were from urban areas (57%) compared to rural areas. Majority (61.2%) were older than 15 years old while 38.2% of cases were younger than 15 years old (Table 2). The mean, median, and standard deviation of age according to their sex were 26.03, 17.50 and 19.73 for males and 23.89, 20.00, 16.56 for females, respectively. According to educational status, 38.2% of cases were illiterate or had no formal education, 34.4% had elementary level education, and 27.3% had middle or higher levels of education (Table 2). As to occupational frequency, shepherds (31.5%), farmers (21.2%), and students (26.9%) ranked the highest while the remaining were registered as other (21.2%).

**Table 2.** Distribution of scorpion sting outcomes according to socio-demographic factors.

| Characteristics |  | Total |  | Outcome |  |  |  |  |  |
| --- | --- | --- | --- | --- | --- | --- | --- | --- | --- |
|  |  |  |  | Recovered |  | Died |  | p-value | OR (95% CI) |
|  |  |  |  | N | % | N | % |  |  |
| Age | <15 years | 63 | 38.2 | 58 | 92.1 | 5 | 7.9 | 0.06 | 2.845 (0.656-12.343) |
|  | ≥15 years | 102 | 61.8 | 99 | 97.1 | 3 | 2.9 | --- | 1 |
| Sex | Male | 80 | 48.5 | 76 | 95.0 | 4 | 5.0 | 0.930 | 1.066 (0.257-4.413) |
|  | Female | 85 | 51.5 | 81 | 95.3 | 4 | 4.7 | --- | 1 |
| Level of Education | No formal education | 63 | 38.2 | 60 | 95.2 | 3 | 4.8 | 0.08 | 1.075 (0.172-6.712) |
|  | Elementary | 57 | 34.4 | 54 | 94.7 | 3 | 5.3 | 0.09 | 1.194 (0.191-7.472) |
|  | Middle or above | 45 | 27.3 | 43 | 95.6 | 2 | 4.4 | --- | 1 |
| Occupation | Farmer | 35 | 21.2 | 34 | 97.1 | 1 | 2.9 | --- | 1 |
|  | Shepard | 52 | 31.5 | 48 | 92.3 | 4 | 7.7 | 0.361 | 2.833 (0.303-26.479) |
|  | Student | 43 | 26.9 | 43 | 100 | 0 | 0.0 | 0.793 | 1.360 (0.136-13.554) |
| Religion | Christian | 45 | 27.3 | 70 | 94.6 | 4 | 5.4 | 0.764 | 1.243 (0.300-5.148) |
|  | Muslim | 120 | 72.7 | 87 | 95.6 | 4 | 4.4 | --- | 1 |

Prior history of scorpion sting was found in 34.5% of the cases while patient experiencing their first sting was found in 65.5% of the cases. The black scorpion (38.8%) was the predominantly identified scorpion followed by yellow/red/white (30.3%) with the rest declared as unknown (31.5%) (Table 3). The site of sting was mostly located on the foot (64.8%) followed by hand (31.5%). Concerning the time of the scorpion sting incident, 62.4% of the sting occurred at night-time compared to 37.6% during the daytime. Indoor stings (56.4%) were slightly higher compared to outdoor stings (43.6%) (Table 3). Of all admitted patients, eight were deceased; thus, the case fatality rate was 4.8%. The fatality outcome was largely associated with class III scorpion sting (p = 0.088).

**Table 3.**
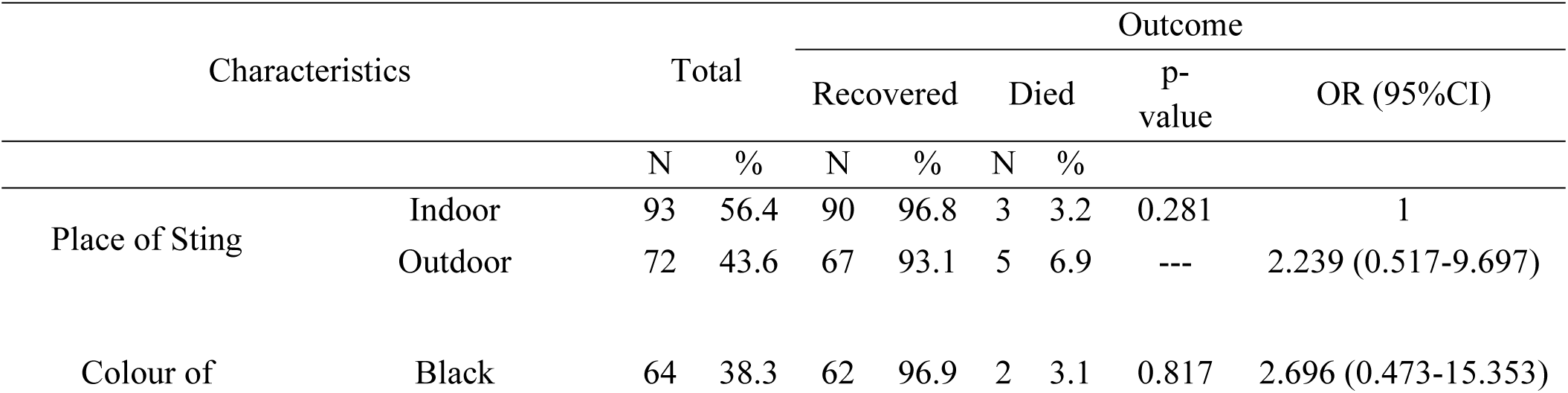
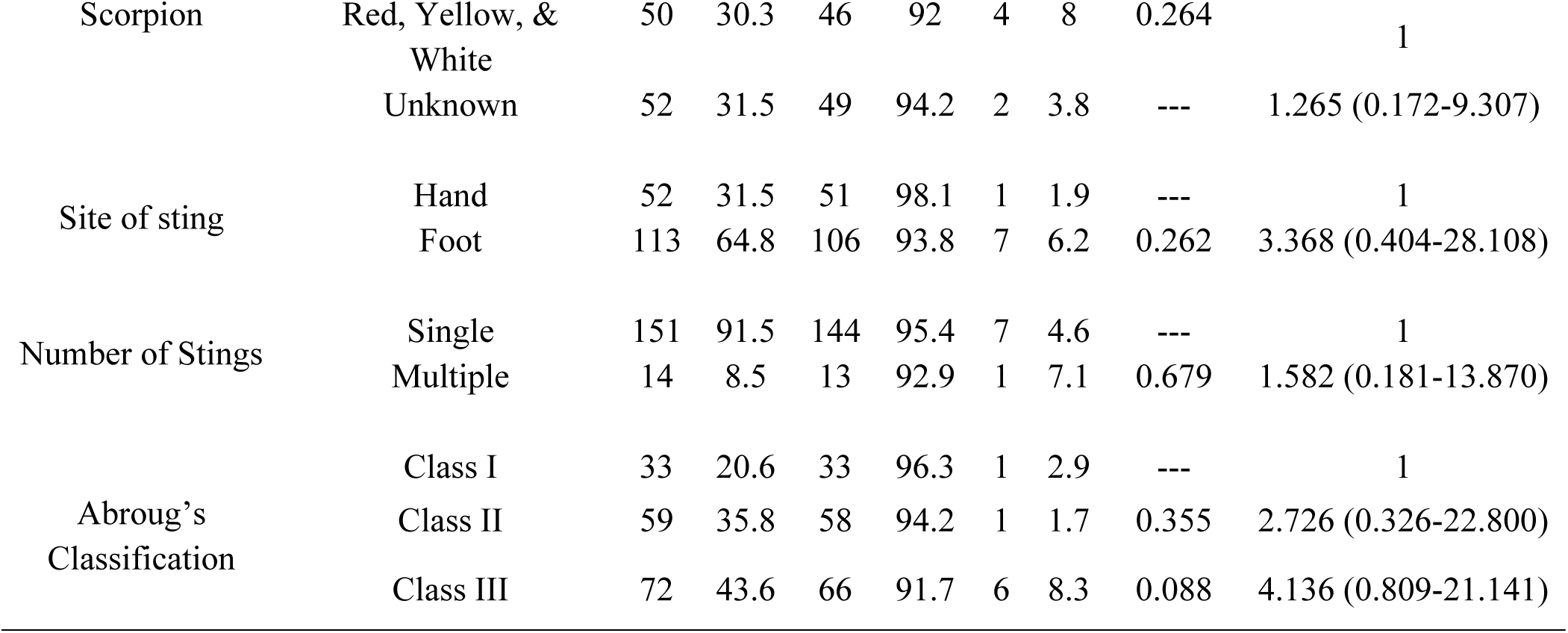
Distribution of scorpion sting outcomes according to scorpion characteristics and Abroug’s classification.

The most common clinical manifestations were localized pain (70.3%), sweating (56.4%), vomiting (33.9%), paraesthesia (29.2%), relative bradycardia (24.8%), restlessness (24.2%), fever (19.4%), and shock (3.6%) (Table 4). Patient subjects less than 15 years old (p = 0.006, OR: 2.845; 95% CI: 0.656-12.343) and those who had no formal education (p = 0.008, OR: 1.075 (0.172-6.712) showed a possible predisposing factor to the severity of symptom (Table 1). Neither the number of stings, the colour of the scorpion, nor the sex of the victim were found to be significant risk factors for severity of symptoms (Table 3). According to Abroug’s classification, 43.6% cases had class three, 35.8% cases had class two, and 20.6% cases had class one (Table 3). Children under five years of age held class III scorpion stings at presentation in 75% of the cases.

**Table 4.** Distribution of scorpion sting envenomation symptoms compared to socio-demographic factors.

| Signs and Symptoms | Sex (%) |  | Age in years (%) |  |  |  | Total (%) |
| --- | --- | --- | --- | --- | --- | --- | --- |
|  | Male | Female | <5 | 5-14 | 15-49 | 50+ |  |
| Fever | 20.0 | 18.8 | 16.7 | 23.5 | 19.0 | 13.0 | 19.4 |
| Elevation of Blood Pressure | 1.3 | 4.7 | 0.0 | 3.9 | 3.8 | 0.0 | 3.0 |
| Relative bradycardia | 20.0 | 29.4 | 50.0 | 29.4 | 20.3 | 17.4 | 24.8 |
| Vomiting | 30.0 | 37.6 | 58.3 | 47.1 | 25.3 | 21.7 | 33.9 |
| Sweating | 60.0 | 52.9 | 83.3 | 66.7 | 49.4 | 43.5 | 56.4 |
| Shock | 3.8 | 3.5 | 16.7 | 3.9 | 1.3 | 4.3 | 3.6 |
| Semi-consciousness | 2.5 | 5.9 | 16.7 | 3.9 | 2.5 | 4.3 | 4.2 |
| Restlessness | 21.3 | 27.1 | 33.3 | 35.3 | 16.5 | 21.7 | 24.2 |
| Convulsion | 1.3 | 1.2 | 16.7 | 0.0 | 0.0 | 0.0 | 1.2 |
| Hypertonic muscles | 1.3 | 3.5 | 0.0 | 2.0 | 3.8 | 0.0 | 2.4 |
| Cyanosis | 0.0 | 1.2 | 0.0 | 0.0 | 1.3 | 0.0 | 0.6 |
| Drooling | 17.5 | 18.8 | 66.7 | 19.6 | 7.6 | 26.1 | 18.2 |
| Paresthesia | 20.0 | 37.6 | 16.7 | 25.5 | 27.8 | 47.8 | 29.1 |
| Dyspepsia | 2.5 | 5.9 | 8.3 | 7.8 | 2.5 | 0.0 | 4.2 |
| Pulmonary edema | 0.0 | 1.2 | 0.0 | 2.0 | 0.0 | 0.0 | 6.0 |
| Generalized pain | 28.8 | 24.7 | 33.3 | 27.5 | 20.3 | 43.5 | 26.7 |
| Localized pain | 66.3 | 74.1 | 50.0 | 70.6 | 78. | 52.2 | 70.3 |

Of all admitted patients, cases with mild symptoms (class I) were managed with local anaesthesia (84.8%) and oral analgesics (41.4%). Patients experiencing generalized pain, restlessness, and convulsions were managed with diazepam (23.6%), with a minority of patients requiring IV fluids (1.9%) and antibiotics (1.2%). Cases with moderate to severe symptoms (class II-III) were managed with Nifedipine and/or SAV (15.2%) (Table 5).

**Table 5.** Distribution of treatment options based on Abroug’s classification.

| Treatment | Class I (%) | Class II (%) | Class III (%) | Total (%) |
| --- | --- | --- | --- | --- |
| Local Anesthesia | 97.1 | 84.7 | 79.2 | 84.8 |
| Oral Analgesics | 14.7 | 40.7 | 54.2 | 41.2 |
| Diazepam | 2.9 | 22.0 | 34.7 | 23.6 |
| IV Fluids | 0.0 | 5.1 | 20.8 | 10.9 |
| Antibiotics | 0.0 | 0.0 | 2.8 | 1.2 |
| Other | 2.9 | 16.9 | 19.4 | 15.2 |

Scorpion sting cases were being admitted throughout the year, the highest case count occurred during the summer season (93%) with September and October being the leading months (Figure 3). Within the summer season, 94.8% of patient cases recovered with seven fatalities during this period.

**Figure 3.**
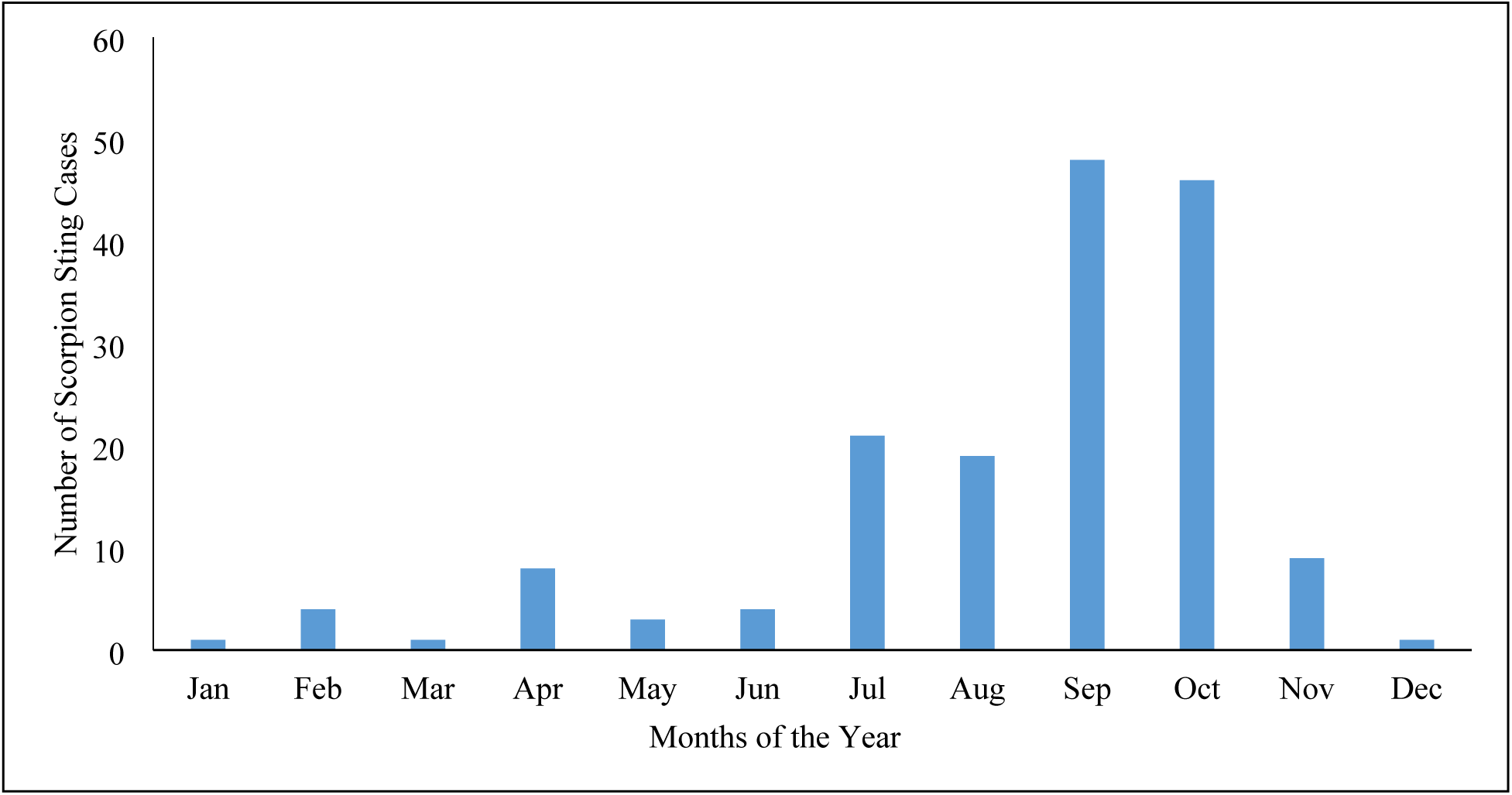
Total number of scorpion sting cases within western lowlands of Eritrea.

## Discussion

Although medical science has advanced in recent decades, scorpion envenomation has largely been ignored due to unknown prevalence of scorpion stings and a high number of low-income victims.(2) Majority of scorpion sting accidents occur in villages in tropical or subtropical regions where the victim do not fully appreciate the seriousness of the medical threat. Thus, victims rely more on traditional healers and remain unregistered in a healthcare setting.(3) In this aspect, to reduce the incidence of scorpion stings and their complications, the 4^th^ International Conference on Snakebites and Scorpion Stings in Dakar stressed the need to develop better recording and reporting systems and optimize the gathering of data on scorpion injuries.(2) The western lowlands of Eritrea have shown to be a conducive area for scorpions with a recent surge of scorpion stings cases within the region, inciting the need for this study. This study is a descriptive, cross-sectional study of scorpionism within the western lowlands of Eritrea with study setting based within the Tesseney Community Hospital. The Tesseney community hospital is the only hospital that receives scorpion sting cases within the region, other corresponding hospital have no similar reports.

Our study held roughly an equal male-to-female ratio (0.94:1). This finding is in contrast to other studies that report higher male-to-female ratio within the Middle East and North Africa (MENA) region where deadly scorpion stings are most common.(2,4,21) Male-to-female ratios differ depending on the study design and region; some studies report higher male-to-females ratios or vice versa.(2,21) These different findings can likely be attributed to the differing social and demographics factors rather than climate and geographical factors. Age groups older than 15 years old were more frequently affected by scorpion stings (61.8%) compared to age groups less than 15 years old (38.2%). The Gash-Barka region is also commonly known as the “bread basket of Eritrea” with the majority of the older male population employed in agriculture or animal herding. Scorpion sting victims older than 15 years old usually performed work outdoors (e.g., shepherd or farmer) and were more likely to be exposed to a scorpion habitat to be stung. Our results report most victims were from an urban (57%) compared to a rural setting. The literature often reports scorpion sting victims typically originate from rural compared to urban areas which contrasts with our findings. Due to the dependence on subsidence agriculture and low socio-economic structure across the Gash-Barka region in which people living in urban areas were using firewood for cooking from rural areas may be attributed to increased number. Beside the criteria classifying rural and urban areas is not definitive. Thus, accurate classification of patient’s place of residence was not possible during the study.

Previous studies report that sting sites are commonly located on the extremities compared to the head or trunk.(4,11,12) Our results showed that 96.2% of sting sites were located in the extremities with a majority of sting sites on the foot (64.8%) compared to the hand (31.5%). Scorpion hiding in shoes left empty overnight or in pockets of loose clothing or trousers are common cause for being stung on the hand or foot.(5) Furthermore, wearing sandals instead of close-toed shoes, working in unprotected fields, and children running barefoot are common occurrence within the Gash Barka region due to the predominate agriculture lifestyle and low socio-economic structure.(4) Our study reports night-time stings (62.4%) were more common compared to daytime stings which corresponds with previous studies.(2,8) An explanation for higher night-time stings may be attributed to residents collecting firewood and storing the firewood inside their residential compound. Firewood is a typical scorpion habitat within the region. As nocturnal animals, scorpions would leave their habitat at night to hunt and may come in contact with a foot or hand of person sleeping on the floor or elevated bed.

Previous studies have shown the prevalence of scorpion sting accidents are more common during the summer season than the winter/autumn season.(2,4) Warm temperature and sunny weather are conditions that leads to higher number of envenomation accidents.(2) Our results aligned with the majority of literature findings; the western lowlands of Eritrea had the highest scorpion sting incidence during the summer period (93%) with September and October being the leading months. The MENA region also report similar conditions with Morocco having the highest scorpion sting incidence between June and September, Saudia Araba between May and September, and Iran between April and October.(4) Our findings showed that black-coloured scorpions (38.3%) were the highest report count followed by yellow/red/white (30.3%), with the remaining cases reported as undetermined (30.9%). A study in Morocco showed similar results with the most common scorpion colour being yellow (41.7%) and black (27.6%) with the remaining being unknown (30.7%).(9) A epidemiological study done in south-eastern Turkey also showed that black and yellow coloured scorpions were predominately identified within their study.(4)

The clinical features of scorpion sting often depend on the subspecies of scorpion, lethality of venom, venom amount injected, age and feeding characteristics of the scorpion as well as seasonal conditions.(4,8) Different species of scorpions have different types of venom which may cause local as well as systemic effects in the first few hours.(1,4,7,9–11,22,24) Systemic manifestations (cardiovascular toxicity, respiratory failure, hypertension and hypotension, etc) are observed at various sites throughout the body and appear within the first 12-24 hours following immediate local effects around the sting site.(4) Within our study, the most common clinical manifestations were localized pain (70.3%), sweating (56.4%), and vomiting (33.9%). Class II-III manifestations were mainly observed amongst age groups less than 15 years old with cardiac rhythm disorder, shock, convulsions, and semi-consciousness being the most common manifestations. In terms of cardiac rhythm disorder, bradycardia was more commonly observed compared to tachycardia. Overall, these findings align with previous studies that report the severity of envenomation was found higher in children compared to adults with related mortality rate.(9,13) Age groups less than 15 years old passing away from class II-III manifestations contributed most to the fatality rate (4.8%), likely due to cardiac dysfunction or pulmonary edema.(4) Patient less than 15 years old (p = 0.006) and had an no formal education (p = 0.008) showed a possible predisposing risk factor for the severity of symptom or death. Within the literature, children are often reported with higher severity of scorpion envenomation symptoms, likely attributed to their smaller body size.(9) With a smaller body mass, venom could diffuse and act more rapidly on the autonomic nervous system, thus have a higher likelihood of systemic involvement in which positions an individual to a higher risk of death.

Our results exhibited an unusually high class III envenoming symptoms (43.6%) compared to similar studies in the literature.(1,22) This was largely due to the villager’s mindset towards arachnid bites or stings. Upon a scorpion sting accident, adult victims (greater than 15 years old) would likely stay at home if they feel mild symptoms, relieving their pain by cleaning the sting site using traditional home remedies. Adult victims would likely seek medication attention only if symptoms from the scorpion sting felt moderate to severe. This may take as long as day or two after the incident had occurred. Children scorpion sting victims (less than 15 years old), however, were immediately admitted to a hospital because pain was intolerable. For adults, the longer elapsed time increased the probability of developing systemic effects and experiencing an “autonomic storm”, thus classifying the patient symptoms as class III. If adult patients recognized the severity of the medical threat and were hospitalized early-on, medication management and monitoring would likely prevent systemic effects.

In terms of medication management, Nifedipine and/or SAV were given in replacement of prazosin for cases presenting with class III symptoms. Nifedipine is often used to treat high blood pressure and is considered a 2^nd^ line treatment with subpar results compared to prazosin. Only 15.2% of cases received either Nifedipine and/or SAV to relieve class III symptoms. Ampicillin or ceftriaxone were often used as a prophylaxis for critical patients who appeared septic and already possessed a catheter. Diazepam was only administered when SAV was not given. Diazepam was used as a sedative for patients experiencing convulsions, shock, and/or restlessness. Within the literature, in cases where severe pulmonary edema, neurotoxicity, and circulatory failure are present, oxygen, sublingual nifedipine, digoxin, furosemide, aminophylline, dopamine, vitamin K, and fresh frozen plasma may be given.(4,12,16) Phenobarbital and dexamethasone may be administered for 48 hours in cases with convulsions.(4,12,16)

## Limitation of the Study

Our study is a prospective study with data limited from a single hospital within the Gash-Barka region. Within this region, victims who suffer scorpion stings may not have registered into a healthcare setting, thus did not have their experience documented. There were also limitations to accurate data recording include identification of scorpion colour and definitive rural vs urban criteria when registering patients. Follow-up after discharging patients also was not performed. Thus, fatalities at home days after discharge were often heard via word-of-mouth and were not included in the study. During the study period, medication efficacy was difficult to determine as SAV stocks ran-out and prazosin therapy was also not available due to budget constraints.

## Conclusion

Our results indicate the main clinical features of scorpion stings were localized pain, sweating, and vomiting. Majority of scorpion envenomation victims were older individuals working in agriculture and unaware to a typical scorpion habitat while performing daily activities. The unusual high level of class III symptoms is likely a consequence of the villager mindset toward arachnid stings, thus most cases had immediate effects around the sting site with mild, rarely fatal, systemic involvement. Although the scorpion sting burden is higher in adults compared to children, our study found that the severity of scorpion envenomation is greater in children compared to adults. Envenomation severity was indicated by the high frequency of class III symptoms found in children, such as cardiac rhythm disorder and convulsions, as well as related fatality rate.

## Recommendation

Preventive educational programs targeting at-risk groups are instrumental to prevent scorpion sting accidents and fatalities in resource-limited areas. Enjehay, a small agriculture-based village in the Gash-Barka region, was recently given a prevention health program due to their alarming high scorpion sting case count compared to their small population size. The program advised to wear close-toed shoes, use mosquito nets, and to place firewood outside the residential compound (away from beds and children) as firewood was a common scorpion habitat. Improved awareness of scorpion sting dangers and increased care-seeking behaviour to a health centre compared to using traditional healing practices was observed by the Tesseney healthcare staff, encouraging broader use of the preventive program. It is also suggested that further qualitative and quantitative studies be performed to investigate the awareness of scorpion stings dangers amongst at-risk population groups (i.e., adolescent and children) as well as typical activities that incite a scorpion sting accident. Investigating healthcare workers care of envenomed patients and capacity to distinguish classes of severity during peak season (i.e., September and October) would also be beneficial to determine competency on efficient drug administration in a resource-limited area.

Equipping healthcare facilities with essential drugs (i.e., prazosin therapy) and educating medical professionals on management of complicated cases, particularly paediatric victims, by introducing updated guideline for treatment are essential. Education programs focused on mother-child and adolescent care on precautionary measures needed to prevent scorpion stings are opportunities to reduce probability of scorpion sting accidents.

Surveys to determine the typical habitats of scorpions, seasonal variation of scorpion sting prevalence, and identifying common scorpion species in scorpion sting prevalent regions are important factors to consider when building effective preventative measures. Since most of the stings are at night, mosquito nets can protect scorpion stings. In endemic areas of venomous stings, clothing, beddings, shoes, and packages should be vigorously shaken out and checked for scorpions without blindly putting hands.

Pesticides (i.e., organophosphates, pyrethrums, and chlorinated hydrocarbons) are also known to kill scorpions.

## Data Availability

Data will be available from the hospital registry department.

## Declarations

### Ethical approval

Ethical approval was obtained from zonal branch of Ministry of Health Research and Ethical Approval Committee and a written informed consent obtained from the patient to participate in this report and publish.

### Fund

The research had no any source of fund

### Consent

A written informed consent was taken from the patients and data confidentiality was secure.

### Author’s contribution

**Conceptualization:** Okbu Frezgi, Araia Berhane, Henok Tekie, Adiam Gebreyohannes.

**Methodology:** Okbu Frezgi, Araia Berhane, Hagos Ahmed, Ghide Ghebrewelde, Henok Tekie, Tsegezab Kiflezgi, Abdelaziz Mohamedsied, Yonas Tekie, Abel Alem.

**Investigation:** Okbu Frezgi, Ghide Ghebrewelde, Henok Tekie, Tsegezab Kiflezgi, Abdelaziz Mohamedsied, Yonas Tekie, Abel Alem.

**Formal analysis:** Okbu Frezgi, Araia Berhane, Hagos Ahmed, Henok Tekie.

**Supervision:** Araia Berhane

**Resources:** Okbu Frezgi,

**Validation: \*\*\***

**Visualization:** Tewaldemedhine Gebrejesus,

**Writing – original draft:** Okbu Frezgi

**Writing – review & editing:** Okbu Frezgi, Araia Berhane, Tewaldemedhine Gebrejesus.

## Abbreviations

(MENA): Middle East and North Africa
(SAV): Scorpion Antivenom
(ED): Emergency Department
(WHO): World Health Organization

## References

1. Alkahlout BH, Abid MM, Kasim MM, Haneef SM. Epidemiological review of scorpion stings in Qatar. Saudi Med J. 2015;36(7):851–5.

2. Khatony A, Abdi A, Fatahpour T, Towhidi F. The epidemiology of scorpion stings in tropical areas of Kermanshah province, Iran, during 2008 and 2009. J Venom Anim Toxins Incl Trop Dis [Internet]. 2015;21(45). Available from: 10.1186/s40409-015-0045-4

3. Bawaskar HS, Bawaskar PH. Management of snake bite and scorpion sting. Vol. 60, Quarterly Medical Review. Mumbai: Raptakos, Brett & Co. Ltd.; 2009.

4. Yılmaz F, Arslan ED, Demir A, Kavalci C, Durdu T, Yılmaz MS, et al. Epidemiologic and clinical characteristics and outcomes of scorpion sting in the southeastern region of Turkey. Ulus Travma Acil Cerr Derg. 2013;19(5):417–22.

5. Bawaskar HS, Bawaskar PH. Scorpion sting: update. Japi. 2012;60:46–55.

6. Chippaux J-P. Emerging options for the management of scorpion stings. Drug Des Devel Ther. 2012;6:165–73.

7. Pradesh U, Bansal A, Bansal AK, Kumar A. Clinical profile of scorpion sting from north Uttar Pradesh, India. Int J Med Sci Public Heal. 2015;4(1):134–7.

8. Ali NOM, Ali NOM. Scorpion sting in different regions of Sudan: epidemiological and clinical survey among university students. Int J Bioinforma Biomed Eng. 2015;1(2):147–52.

9. Abourazzak S, Achour S, El Arqam L, Atmani S, Chaouki S, Semlali I, et al. Epidemiological and clinical characteristics of scorpion stings in children in Fez, Morocco. J Venom Anim Toxins incl Trop Dis. 2009;15(2):255–67.

10. Riaño-umbarila L, Ilse VG, Ledezma-candanoza LM, Olamendi-portugal T, Remi E. Generation of a broadly cross-neutralizing antibody fragment against several mexican scorpion venoms. Toxins (Basel). 2019;11(32):1–17.

11. Soren C, Nageswara Rao K. Clinical profile of scorpion sting envenomation in children. Int J Contemp Pediatr. 2016;3(3).

12. Rao KS, Medicine G, Surgeon C. Clinical study of scorpion sting envenomation - clinical profile, complications, treatment and effect of early treatment with prazocin in preventing the complications. Int J Sci Res. 2021;10(9):7–10.

13. Singhal A, Mannan R, Rampal U. Epidemiology, clinical presentation and final outcome of patients with scorpion bite. J Clin Diagnostic Res. 2009;3(3):1523–8.

14. Bawaskar HS, Bawaskar PH. Efficacy and safety of scorpion antivenom plus prazosin compared with prazosin alone for venomous scorpion (Mesobuthus tamulus) sting: randomised open label clinical trial. BMJ. 2011;

15. Murthy KRK. On scorpion envenoming syndrome: problems of medical ethics and accountability in medical research in India. J Venom Anim Toxins. 2002;8(1):3–17.

16. Agrawal A, Kumar A, Consul S, Yadav A. Scorpion bite, a sting to the heart! Indian J Crit Care Med. 2015;19(4):233–6.

17. LoVecchio F. Scorpion envenomation causing autonomic dysfunction (North Africa, Middle East, Asia, South America, and the Republic of Trinidad and Tobago). 2023.

18. Barish RA, Arnold T. Scorpion Stings. In: Merck Manual Consumer Version. 2022.

19. Klotz SA, Yates S, Smith SL, Dudley Jr S, Schmidt JO, Shirazi FM. Scorpion Stings and Antivenom Use in Arizona -. Am J Med. 2021;134(8):1034–8.

20. Shamoon Z, Peterfy RJ, Hammoud S, Khazaeni B. Scorpion Toxicity. In: StatPearls. StatPearls Publishing LLC; 2022.

21. Kassiri H, Mahijan NM, Hasanvand Z, Shemshad M, Shemshad K, Control V, et al. Epidemiological survey on scorpion sting envenomation in south-west, Iran. Zahedan J Reserach Med Sci. 2012;14(8):80–3.

22. Duman A, Dagli B, Akoz A, Avcil M, Orun S, Turkdoğan KA. Importance of antivenom in management of scorpion envenomation with epidemiologic and clinical characteristics. Biomed Res. 2017;28(8):3763–8.

23. Gupta BD, Parakh M, Purohit A. Management of scorpion sting: prazosin or dobutamine. J Trop Pediatr. 2010;56(2):115–8.

24. Bhadani UK, Tripathi M, Sharma S, Pandey R. Scorpion sting envenomation presenting with pulmonary edema in adults: a report of seven cases from Nepal. Indian J Med Sci. 2006;60(1):20–2.

